# Predictors of COVID-19 vaccine uptake in healthcare workers: a cross-sectional study in Greece

**DOI:** 10.1101/2021.09.14.21263300

**Authors:** Petros Galanis, Ioannis Moisoglou, Irene Vraka, Olga Siskou, Olympia Konstantakopoulou, Aglaia Katsiroumpa, Daphne Kaitelidou

## Abstract

**Background:** The role of healthcare workers (HCWs) in the general public health is crucial and their decision to vaccinate against the COVID-19 can have a positive impact on the general population facilitating widespread COVID-19 vaccine uptake.

**Objective:** To estimate the uptake of a COVID-19 vaccine in HCWs and to expand our knowledge regarding the predictors of COVID-19 vaccine uptake.

**Methods:** An on-line cross-sectional study was conducted in Greece during August 2021. We collected socio-demographic data of HCWs and we measured attitudes towards vaccination and COVID-19, knowledge and trust. We used a convenience sample since we distributed the questionnaire through social media and e-mails.

**Results:** Study population included 855 HCWs. The majority of HCWs were vaccinated against the COVID-19 (91.5%). According to multivariate analysis, females, HCWs without a previous COVID-19 diagnosis, and HCWs with previous seasonal influenza vaccination history had a greater probability to take a COVID-19 vaccine. Also, increased self-perceived knowledge regarding COVID-19 and increased trust in COVID-19 vaccines and government regarding the information about the COVID-19 vaccines were associated with COVID-19 vaccine uptake. On the other hand, HCWs with more concerns about the side-effects of COVID-19 vaccination were more reluctant to take a COVID-19 vaccine.

**Conclusions:** Our study provides a timely assessment of COVID-19 vaccination status among HCWs and identifies specific factors associated with COVID-19 vaccine uptake. By understanding these factors, policy makers and scientists will be able to develop novel strategies to improve COVID-19 vaccine uptake among HCWs.

## Introduction

Several vaccines are effective in preventing the severe acute respiratory syndrome coronavirus 2 (SARS-CoV-2) and they are being used throughout the world (Baden et al., 2021; Logunov et al., 2021; Polack et al., 2020; Wu et al., 2021). The widespread use of COVID-19 vaccines is critical to control the COVID-19 pandemic, but several reasons could delay or decline COVID-19 vaccine uptake. According to a systematic review, the most important reasons for decline of a COVID-19 vaccine are concerns about the safety and effectiveness of COVID-19 vaccines, medical conditions, religious and ethical reasons, pregnancy, fertility, limited knowledge, and previous COVID-19 diagnosis (Galanis et al., 2021). COVID-19 vaccine hesitancy limits general population protection from SARS-CoV-2. The situation is getting worse in case of healthcare workers (HCWs), since they are at higher risk of exposure and transmission of the SARS-CoV-2, and they could put themselves, co-workers and patients at risk.

To our knowledge, literature regarding COVID-19 vaccine uptake among HCWs is still poor since six studies have been conducted in this field and only one in Europe (Barry et al., 2021; Gharpure et al., 2021; Martin et al., 2021; Pacella-LaBarbara et al., 2021; Schrading et al., 2021; Xu et al., 2021). The results showed that the uptake of a COVID-19 vaccine among HCWs is different ranging from 33.3% in Kingdom of Saudi Arabia (Barry et al., 2021) and 64.5% in United Kingdom (Martin et al., 2021), to 86.2% in China (Xu et al., 2021) and 94.5% in the USA (Schrading et al., 2021). Moreover, HCWs intention to accept COVID-19 vaccination is moderate (63.5%) according to a meta-analysis included 24 studies and 39,617 participants worldwide (Galanis et al., 2020). Several socio-demographic factors increase HCWs’ uptake of a COVID-19 vaccine, e.g. male gender, older age, higher educational level, white race, etc. (Galanis et al., 2021).

To date, only one study on the actual acceptance of a COVID-19 vaccine in HCWs in Europe is reported (Martin et al., 2021). Moreover, research until now focus only on socio-demographic determinants of COVID-19 vaccine in HCWs. Thus, we aimed to estimate the uptake of a COVID-19 vaccine in a sample of HCWs in Greece and to expand our knowledge regarding the predictors of COVID-19 vaccine uptake.

## Methods

### Study design and participants

An on-line cross-sectional study was conducted in Greece during August 2021. From January 2021 until the time of study, a free COVID-19 vaccine was offered from Greek government to all HCWs throughout the country. The vaccine was taken on a voluntary basis and was offered irrespective of past history of COVID-19. We used google forms to create an anonymous version of the study questionnaire. We used a convenience sample since we distributed the questionnaire through social media and e-mails. The on-line questionnaire was accompanied by a detailed explanation of the study aim and design, and HCWs provided informed consent to participate anonymously in the study. All HCWs over 18 years old were allowed to participate in the study. Given the wide range of COVID-19 vaccine uptake among the HCWs through the studies, we considered a prevalence of 50% to estimate the largest sample size for our study. Thus, considering prevalence of COVID-19 vaccine uptake as 50%, precision level as 5%, and confidence level as 95%, we calculated a minimum sample size of 385 HCWs. We decided to increase substantially the sample size to minimize random error. The Ethics Committee of Department of Nursing, National and Kapodistrian University of Athens approved the study protocol (reference number; 370, 02-09-2021).

### Questionnaire

We collected the following socio-demographic data of HCWs: gender, age, marital status, under-age children, educational level, profession, years of experience, self-perceived financial status, self-perceived health status, chronic disease, previous COVID-19 diagnosis, family/friends with previous COVID-19 diagnosis, living with elderly people or vulnerable groups during the COVID-19 pandemic, and providing care to COVID-19 patients. Financial status and self-perceived health status were measured in a five-point Likert scale from 0 to 4 (0=“very poor”, 1=“poor”, 2=“moderate”, 3=“good”, and 4=“very good”).

Regarding vaccination, we measured seasonal influenza vaccination in 2020 and COVID-19 vaccination with “yes/no” answers. Moreover, we recorded possible reasons for decline of COVID-19 vaccination, e.g. concerns about the safety and effectiveness of COVID-19 vaccines, fear for side-effects, religious reasons, pregnancy, previous COVID-19 diagnosis, etc.

Also, we measured self-perceived severity of COVID-19, self-perceived knowledge regarding COVID-19 and COVID-19 vaccines, concerns about the side-effects of COVID-19 vaccination, trust in COVID-19 vaccines, and trust in the government, scientists and family doctors regarding the information about the COVID-19 vaccines on a scale from 0 to 10 with higher values indicate higher levels of self-perceived severity of COVID-19, knowledge, concerns, and trust.

### Statistical analysis

We used numbers (percentages) to present categorical variables and mean (standard deviation) to present continuous variables. The Kolmogorov-Smirnov test and normal Q-Q plots were applied to test the normality of the distribution of the continuous variables. COVID-19 vaccination was the dependent variable and we defined the outcome as 1 if a HCW took a COVID-19 vaccine. First we performed univariate logistic regression analysis and independent variables with p-values <0.20 were included in a multivariate logistic regression model to eliminate confounding. We applied a backward stepwise model and we calculated adjusted odds ratios (OR), 95% confidence intervals (CI) and p-values. In multivariate logistic regression model, p-values<0.05 were considered significant. All tests of statistical significance were two-tailed. Statistical analysis was performed with the Statistical Package for Social Sciences software (IBM Corp. Released 2012. IBM SPSS Statistics for Windows, Version 21.0. Armonk, NY: IBM Corp.).

## Results

Study population included 855 HCWs. Detailed socio-demographic characteristics of HCWs are shown in Table 1. Mean age of HCWs was 40.9 years and mean years of clinical experience were 14.4. Among our HCWs, 80.7% were females, 48.9% had a MSc/PhD degree, 45.3% were nurses, and 20.1% have suffered from a chronic disease. Regarding the COVID-19, 10.8% of HCWs were diagnosed with COVID-19, 58.8% had family/friends with a previous COVID-19 diagnosis, and 49.8% provided care to COVID-19 patients. Most of the HCWs considered their financial status as moderate/good (83.6%) and their health status as good/very good (81.9%).

**Table 1.** Socio-demographic characteristics of healthcare workers.

| **Characteristics** | **N** | **%** |
| --- | --- | --- |
| Gender |  |  |
| Females | 714 | 80.7 |
| Males | 171 | 19.3 |
| Age (years)<sup>a</sup> | 40.9 | 9.9 |
| Marital status |  |  |
| Singles | 254 | 28.7 |
| Married | 565 | 63.8 |
| Widowed | 61 | 6.9 |
| Divorced | 5 | 0.6 |
| Children <18 years old |  |  |
| No | 398 | 45.0 |
| Yes | 487 | 55.0 |
| MSc/PhD degree |  |  |
| No | 452 | 51.1 |
| Yes | 433 | 48.9 |
| Profession |  |  |
| Physicians | 220 | 25.2 |
| Nurses | 396 | 45.3 |
| Nurses assistants | 47 | 5.4 |
| Midwives | 16 | 1.8 |
| Paramedics | 73 | 8.4 |
| Administrative staff | 72 | 8.2 |
| Pharmacists | 28 | 3.2 |
| Biochemists | 7 | 0.8 |
| Dentists | 5 | 0.6 |
| Ambulatory staff | 10 | 1.1 |
| Clinical experience (years)<sup>a</sup> | 14.4 | 9.5 |
| Self-perceived financial status |  |  |
| Very poor | 10 | 1.1 |
| Poor | 80 | 9.0 |
| Moderate | 483 | 54.6 |
| Good | 257 | 29.0 |
| Very good | 55 | 6.2 |
| Self-perceived health status |  |  |
| Very poor | 3 | 0.3 |
| Poor | 17 | 1.9 |
| Moderate | 140 | 15.8 |
| Good | 446 | 50.4 |
| Very good | 279 | 31.5 |
| Chronic disease |  |  |
| No | 707 | 79.9 |
| Yes | 178 | 20.1 |
| Previous COVID-19 diagnosis |  |  |
| No | 789 | 89.2 |
| Yes | 96 | 10.8 |
| Family/friends with previous COVID-19 diagnosis |  |  |
| No | 365 | 41.2 |
| Yes | 520 | 58.8 |
| Living with elderly people or vulnerable groups during the COVID-19 pandemic |  |  |
| No | 626 | 70.7 |
| Yes | 259 | 29.3 |
| Providing care to COVID-19 patients |  |  |
| No | 441 | 50.2 |
| Yes | 438 | 49.8 |

<sup>a</sup> mean, standard deviation

Table 2 presents HCWs’ attitudes towards COVID-19 vaccination and pandemic. The majority of HCWs were vaccinated against the COVID-19 (91.5%), while the respective percentage for the seasonal influenza in 2020 was 64.6%. The most important reasons for decline of COVID-19 vaccination were concerns about the safety and effectiveness of COVID-19 vaccines (50%), concerns about the side-effects of COVID-19 vaccines (17.6%), previous COVID-19 diagnosis (12.2%), and females’ effort to get pregnant (9.5%). HCWs reported high levels of knowledge regarding COVID-19 and COVID-19 vaccines and moderate concerns about the side-effects of COVID-19 vaccination. Regarding the information about the COVID-19 vaccines, HCWs showed more trust in family doctors and scientists than in the government.

**Table 2.**
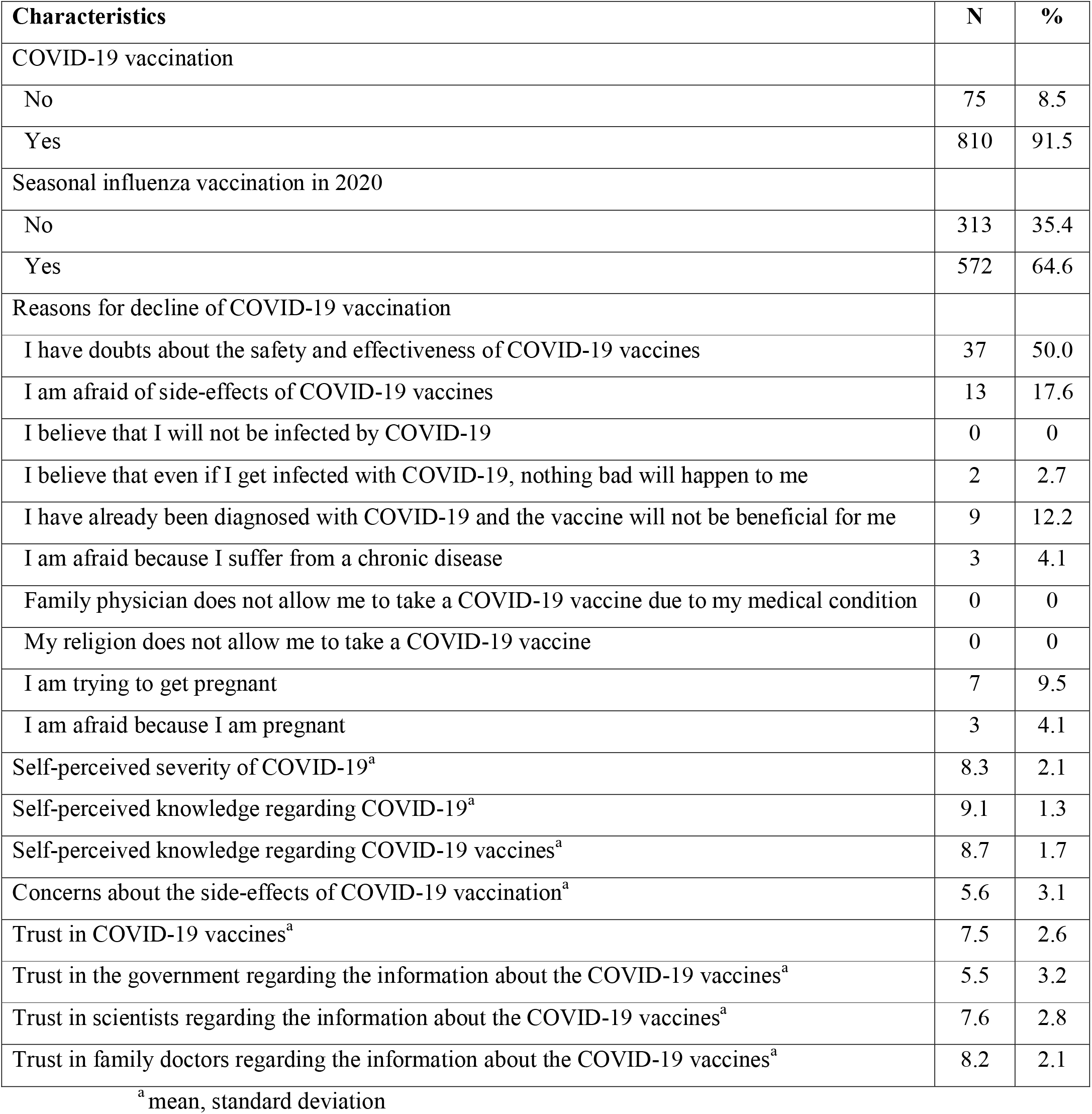
Healthcare workers’ attitudes towards COVID-19 vaccination and pandemic.

| Characteristics | N | % |
| --- | --- | --- |
| COVID-19 vaccination |  |  |
| No | 75 | 8.5 |
| Yes | 810 | 91.5 |
| Seasonal influenza vaccination in 2020 |  |  |
| No | 313 | 35.4 |
| Yes | 572 | 64.6 |
| Reasons for decline of COVID-19 vaccination |  |  |
| I have doubts about the safety and effectiveness of COVID-19 vaccines | 37 | 50.0 |
| I am afraid of side-effects of COVID-19 vaccines | 13 | 17.6 |
| I believe that I will not be infected by COVID-19 | 0 | 0 |
| I believe that even if I get infected with COVID-19, nothing bad will happen to me | 2 | 2.7 |
| I have already been diagnosed with COVID-19 and the vaccine will not be beneficial for me | 9 | 12.2 |
| I am afraid because I suffer from a chronic disease | 3 | 4.1 |
| Family physician does not allow me to take a COVID-19 vaccine due to my medical condition | 0 | 0 |
| My religion does not allow me to take a COVID-19 vaccine | 0 | 0 |
| I am trying to get pregnant | 7 | 9.5 |
| I am afraid because I am pregnant | 3 | 4.1 |
| Self-perceived severity of COVID-19<sup>a</sup> | 8.3 | 2.1 |
| Self-perceived knowledge regarding COVID-19<sup>a</sup> | 9.1 | 1.3 |
| Self-perceived knowledge regarding COVID-19 vaccines<sup>a</sup> | 8.7 | 1.7 |
| Concerns about the side-effects of COVID-19 vaccination<sup>a</sup> | 5.6 | 3.1 |
| Trust in COVID-19 vaccines<sup>a</sup> | 7.5 | 2.6 |
| Trust in the government regarding the information about the COVID-19 vaccines<sup>a</sup> | 5.5 | 3.2 |
| Trust in scientists regarding the information about the COVID-19 vaccines<sup>a</sup> | 7.6 | 2.8 |
| Trust in family doctors regarding the information about the COVID-19 vaccines<sup>a</sup> | 8.2 | 2.1 |

<sup>a</sup> mean, standard deviation

Logistic regression analysis is shown in Table 3. According to multivariate logistic regression analysis, eight variables were related with COVID-19 vaccine uptake in healthcare workers. In particular, females, HCWs without a previous COVID-19 diagnosis, and HCWs with previous seasonal influenza vaccination history had a greater probability to take a COVID-19 vaccine. Increased self-perceived knowledge regarding COVID-19 and increased trust in COVID-19 vaccines and government regarding the information about the COVID-19 vaccines were associated with COVID-19 vaccine uptake. On the other hand, HCWs with more concerns about the side-effects of COVID-19 vaccination were more reluctant to take a COVID-19 vaccine. Moreover, increased self-perceived knowledge regarding COVID-19 vaccines was associated with COVID-19 vaccine hesitancy.

**Table 3.** Univariate and multivariate logistic regression analysis with COVID-19 vaccine uptake in healthcare workers as the dependent variable (reference: COVID-19 vaccine denial).

| Variable | Unadjusted OR (95% CI) | P-value | Adjusted OR (95% CI)<sup>a</sup> | P-value |
| --- | --- | --- | --- | --- |
| Gender (females vs. males) | 1.47 (0.85 – 2.54) | 0.17 | 3.13 (1.27 – 7.70) | 0.01 |
| Age (years) | 1.01 (0.98 – 1.03) | 0.59 | NS |  |
| Marital status (married vs. singles/widowed/divorced) | 1.12 (0.69 – 1.83) | 0.64 | NS |  |
| Children <18 years old (no vs. yes) | 1.71 (1.04 – 2.81) | 0.04 | NS |  |
| MSc/PhD degree (yes vs. no) | 1.32 (0.82 – 2.13) | 0.26 | NS |  |
| Profession |  |  | NS |  |
| Physicians | 4.31 (1.76 – 10.54) | 0.001 |  |  |
| Nurses | 3.18 (1.45 – 7.00) | 0.004 |  |  |
| Administrative staff | 2.16 (0.78 – 5.96) | 0.14 |  |  |
| Paramedics | 2.55 (0.89 – 7.26) | 0.08 |  |  |
| Others | 4.19 (1.23 – 14.32) | 0.02 |  |  |
| Nurses assistants | 1 (reference) |  |  |  |
| Clinical experience | 1.01 (0.98 – 1.04) | 0.54 | NS |  |
| Self-perceived financial status |  |  | NS |  |
| Good/very good | 2.59 (1.31 – 5.13) | 0.006 |  |  |
| Moderate | 2.77 (1.46 – 5.25) | 0.002 |  |  |
| Very poor/poor | 1 (reference) |  |  |  |
| Self-perceived health status |  |  | NS |  |
| Good/very good | 2.82 (0.91 – 8.72) | 0.07 |  |  |
| Moderate | 2.67 (0.77 – 9.26) | 0.12 |  |  |
| Very poor/poor | 1 (reference) |  |  |  |
| Chronic disease (yes vs. no) | 1.01 (0.56 – 1.92) | 0.98 | NS |  |
| COVID-19 disease (no vs. yes) | 2.96 (1.66 – 5.29) | <0.001 | 3.23 (1.33 – 7.86) | 0.01 |
| Family/friends with COVID-19 disease (no vs. yes) | 1.77 (1.06 – 2.97) | 0.03 | NS |  |
| Living with elderly people or vulnerable groups during the COVID-19 pandemic (no vs. yes) | 1.40 (0.85 – 2.30) | 0.18 | NS |  |
| Providing care to COVID-19 patients (no vs. yes) | 1.01 (0.63 – 1.62) | 0.98 | NS |  |
| Seasonal influenza vaccination in 2020 (yes vs. no) | 7.43 (4.24 – 13.01) | <0.001 | 4.57 (2.17 – 9.61) | <0.001 |
| Self-perceived severity of COVID-19 | 1.56 (1.42 – 1.72) | <0.001 | NS |  |
| Self-perceived knowledge regarding COVID-19 | 1.17 (1.01 – 1.35) | 0.04 | 1.42 (1.08 – 1.88) | 0.01 |
| Self-perceived knowledge regarding COVID-19 vaccines | 1.12 (0.99 – 1.26) | 0.09 | 0.67 (0.53 – 0.86) | 0.001 |
| Concerns about the side-effects of COVID-19 vaccination | 0.56 (0.48 – 0.64) | <0.001 | 0.74 (0.62 – 0.88) | 0.001 |
| Trust in COVID-19 vaccines | 1.77 (1.61 – 1.94) | <0.001 | 1.55 (1.31 – 1.82) | <0.001 |
| Trust in the government regarding the information about the COVID-19 vaccines | 1.55 (1.41 – 1.72) | <0.001 | 1.18 (1.02 – 1.37) | 0.03 |
| Trust in scientists regarding the information about the COVID-19 vaccines | 1.52 (1.40 – 1.65) | <0.001 | NS |  |
| Trust in family doctors regarding the information about the COVID-19 vaccines | 1.38 (1.26 – 1.51) | <0.001 | NS |  |

An odds ratio <1 indicates a negative association, while an odds ratio >1 indicates a positive association.
CI: confidence interval; NS: not selected by the backward elimination procedure in the multivariable logistic regression analysis with a significance level set at 0.05; OR: odds ratio
<sup>a</sup> $R^2$ for the final multivariate model was 57%
<sup>b</sup> Due to low number of healthcare workers, we merged the following categories: “very poor” and “poor”; “good” and “very good”

## Discussion

We conducted a study to estimate COVID-19 vaccine uptake in a sample of HCWs in Greece and investigate the predictors of this uptake. A great percentage of our HCWs (91.5%) have been vaccinated against the COVID-19. This percentage appears similar to that found in studies in the USA (94.5%) and China (86.2%) (Schrading et al., 2021; Xu et al., 2021). On the other hand, four studies in the USA, United Kingdom, and Kingdom of Saudi Arabia found lower COVID-19 vaccine uptake from to 33.3% to 79% (Barry et al., 2021; Gharpure et al., 2021; Martin et al., 2021; Pacella-LaBarbara et al., 2021). Data collection time could explain this variability in COVID-19 vaccine uptake since the closer each study was performed to now, the more likely HCWs were to take a COVID-19 vaccine. At the time of our study, COVID-19 vaccination for HCWs was voluntary in Greece but the government was planning a mandatory vaccination program for HCWs and other occupational groups from September 2021. This intention of the Greek government could explain in part the high percentage of COVID-19 vaccine uptake in HWCs in our study.

Our multivariate regression model revealed varied factors were associated with COVID-19 vaccine uptake in HCWs. Specifically, trust in COVID-19 vaccines and less concerns about the side-effects of COVID-19 vaccination were associated with vaccine acceptance. This finding is confirmed by the literature since the main reasons for the decline of COVID-19 vaccination include concerns about the COVID-19 vaccine safety and effectiveness (Schrading et al., 2021; Xu et al., 2021). Thus, policy makers and scientists should provide unvaccinated HCWs with more safety and surveillance data of the COVID-19 vaccines.

Moreover, we found that increased self-perceived knowledge regarding COVID-19 vaccines is associated with COVID-19 vaccine hesitancy. High levels of self-perceived knowledge among HCWs do not mean necessarily adequate knowledge regarding COVID-19 vaccines since many sources of information (e.g. social media, religious leaders, etc.) are spurious and misleading during the COVID-19 pandemic. Detection of fake news is associated with intention to take a COVID-19 vaccine (Montagni et al., 2021). Also, COVID-19 vaccine uptake is higher among individuals that do not use social media as a source of information during the COVID-19 pandemic (Barry et al., 2021). Research indicates that on-line COVID-19 information from most websites is of poor quality and inadequate (Cuan-Baltazar et al., 2020; Fan et al., 2020; Joshi et al., 2020). During the COVID-19 pandemic, a major health crisis misinformation is produced by the media, and the misinformation is obtained by HCWs from the websites. Additionally, information regarding COVID-19 vaccines is of particular interest, since these vaccines are an innovation and new data are constantly emerging. Governments should develop strategies to regulate COVID-19 information on the internet ensuring that websites will provide evidence-based information related to COVID-19 vaccines.

Our findings demonstrate higher COVID-19 vaccine uptake among HCWs with previous seasonal influenza vaccination history. The role of influenza vaccination in the uptake of COVID-19 vaccine in HCWs has not been investigated in other studies but has already been shown to be critical in the intention of HCWs to accept a COVID-19 vaccine (Galanis et al., 2020). Unfortunately, influenza vaccination rate among HCWs is low although is higher than general population and high-risk groups (Blank et al., 2008; La Torre et al., 2011; Sheldenkar et al., 2019; Wang et al., 2018). Refusal of influenza vaccination is evidence of vaccine hesitancy, one of the top ten threats to global health in 2019 according to the World Health Organization (World Health Organization, 2020). The negative attitude of HCWs towards vaccination is already known (Di Martino et al., 2020; Lau et al., 2020; Wilson et al., 2020) and COVID-19 vaccine hesitancy in HCWs is crucial as it can undermine public confidence (MacDonald & Dubé, 2015; Opel et al., 2013). Educational programs and workplace strategies are proven effective to improve influenza vaccination coverage among HCWs and may also serve as a guide to improve COVID-19 vaccine uptake (Black et al., 2018).

Consistent with prior literature (Martin et al., 2021; Pacella-LaBarbara et al., 2021), HCWs with a history of COVID-19 diagnosis were more likely to be unvaccinated. Individuals with a history of COVID-19 diagnosis are likely to feel less risk of being re-infected by the virus and/or have severe consequences. Risk perception is critical to vaccination intention, since as risk perception increases, so does the intention to accept a COVID-19 vaccine (Caserotti et al., 2021). Also, when the risk of contracting COVID-19 is low, intention on taking a COVID-19 vaccine is low (Karlsson et al., 2021).

We found that females had greater COVID-19 vaccine uptake than males. This finding is interesting since it is in contrary to the previous studies (Barry et al., 2021; Martin et al., 2021; Pacella-LaBarbara et al., 2021; Schrading et al., 2021). In general, COVID-19 vaccine hesitancy is more common among females (Gagneux-Brunon et al., 2020; Nzaji MK et al., 2020; Shaw et al., 2021; Unroe et al., 2021; Verger et al., 2021). Our finding may be due to the fact that we now have more knowledge about the safety and effectiveness of COVID-19 vaccines. For instance, the results of recent studies show the effectiveness of vaccines in both pregnant and lactating women (Ciapponi et al., 2021; Garg et al., 2021).

Our study suffers from several limitations. Although our study population was large, we used a convenience sample which is not representative of HCWs in Greece. Additionally, response rate cannot be calculated since we conducted an on-line study. Moreover, vaccine uptake and other information were self-reported and social desirability to bias responses may exist. For instance, some HCWs may have falsely stated that they had received a COVID-19 vaccine. We used an anonymous on-line questionnaire to reduce this bias. Further, we investigated a variety of determinants of COVID-19 vaccine uptake and some of them had not been studied before. However, it is possible that there are other factors affecting COVID-19 vaccination. Future research may consider including other factors which may influence COVID-19 vaccine uptake, e.g. personality traits, social media variables, fake news, conspiracy theories, etc. Finally, as is always the case in cross-sectional studies, no causal relationships between independent variables and COVID-19 vaccine uptake can be established.

## Conclusions

Our study provides a timely assessment of COVID-19 vaccination status among HCWs in Greece and identifies specific factors associated with COVID-19 vaccine uptake. Future work is needed to understand the factors influencing the decision of HCWs to vaccinate against the COVID-19. By understanding these factors, policy makers and scientists will be able to develop novel strategies to improve COVID-19 vaccine uptake among HCWs. The role of HCWs in the general public health is crucial and their decision to vaccinate can have a positive impact on the general population facilitating widespread COVID-19 vaccine uptake.

## Data Availability

Data will be available after reasonable request

